# Genomic characterization of the 2024/2025 Mpox outbreak in Uganda

**DOI:** 10.64898/2026.03.16.26348494

**Authors:** Stephen Kanyerezi, Alisen Ayitewala, Andrew Nsawotebba, Caroline Makoha, Godwin Tusabe, Jupiter Marina Kabahita, Hellen Rosette Oundo, Julius Seruyange, Wilson Tenywa, Stacy Were, Moses Murungi, Valeria Nakintu, Ivan Sserwadda, Harris Onywera, Collins Tanui, Ibrahim Mugerwa, Atek Kagirita, Benard Lubwama, Eilu Roggers Michael, David Patrick Kateete, Morgan Otita, Samuel Giduddu, Daudi Jjingo, Gerald Mboowa, Aloysious Ssemaganda, Susan Nabadda, Sofonias K Tessema, Isaac Ssewanyana

**Affiliations:** Department of National Health Laboratory and Diagnostics Services, Central Public Health Laboratories (NHLDS-CPHL), Ministry of Health, P.O. Box 7272, Kampala, Uganda; The African Center of Excellence in Bioinformatics and Data-Intensive Sciences, the Infectious Diseases Institute, College of Health Sciences, Makerere University, P.O Box 22418, Kampala, Uganda; Department of Immunology and Molecular Biology, School of Biomedical Sciences, College of Health Sciences, Makerere University, P.O Box 7072, Kampala, Uganda; Africa Centers for Disease Control and Prevention (Africa CDC), Addis Ababa, Ethiopia; Amsterdam Institute for Global Health and Development (AIGHD), Department of Global Health, Academic Medical Center, Amsterdam, The Netherlands; Broad Institute of Harvard and MIT, Cambridge, MA 02142, USA; National Public Health Emergency Operations Center-PEOC, Ministry of Health; Department of Global Health Security, Infectious Diseases Institute, Makerere University, Kampala, Uganda; Uganda National Institute of Public Health, Ministry of Health; Infectious Diseases Research Collaboration, Kampala, Uganda

**Author notes:** To whom correspondence should be addressed: Isaac Ssewanyana.

**Keywords:** Mpox, Uganda, Genomic Epidemiology, ARTIC Pipeline, Nextstrain

## Abstract

Mpox has historically been endemic in Central and West Africa, driven by recurrent zoonotic spillover events, but recent outbreaks in East Africa underscore its expanding geographic footprint. Despite this shift, genomic data from East Africa remain limited. We performed genomic characterization of the 2024/2025 Mpox outbreak in Uganda using PCR-confirmed monkeypox virus (MPXV) positive samples (n=511) from 44 districts, all achieving ≥70% genome coverage. To provide regional context, we incorporated 895 publicly available clade Ib MPXV genomes from GISAID, Pathoplexus, and NCBI.

Phylogenetic analysis revealed two major clusters within clade Ib, each subdivided into two subclusters, indicating substantial viral diversification. Most Ugandan sequences clustered within the most genetically diverse subcluster. Additional Ugandan genomes were distributed across other subclusters, indicating co-circulation of multiple lineages. Cluster 1 was dominated by sequences from the Democratic Republic of Congo, while phylogeographic analysis identified multiple cross-border introductions into Uganda.

These findings highlight the role of regional connectivity in shaping MPXV transmission and underscore the importance of integrated genomic surveillance and cross-border data sharing to inform outbreak response in East and Central Africa.

## Introduction

Monkeypox virus (MPXV), a zoonotic double-stranded DNA virus belonging to the genus Orthopoxvirus (family Poxviridae), has re-emerged as a significant global public health threat over the past decade [1]. Historically confined to forested regions of Central and West Africa, MPXV has increasingly been reported in urban and peri-urban settings, reflecting changes in human behavior, population mobility, and ecological interfaces [2], [3]. The unprecedented global outbreak of 2022 highlighted the virus’s capacity for sustained human-to-human transmission, leading to intensified genomic surveillance efforts worldwide and renewed interest in understanding MPXV evolution, transmission dynamics, and adaptation in human populations [4].

Mpox has historically been endemic in forested regions of Central and West Africa, with recurrent zoonotic spillover events [5]. In recent years, however, outbreaks have increasingly been reported in non-endemic regions, including East Africa [6]–[8], underscoring the need for expanded surveillance and genomic characterization across diverse epidemiological settings. Despite the growing number of reported outbreaks, genomic data from these regions remain comparatively limited [2]. This imbalance has constrained comprehensive understanding of MPXV evolutionary trajectories within endemic regions and limited the ability to contextualize global MPXV diversity using African reference genomes. Genomic characterization of outbreaks in these settings is therefore critical, not only for local public health response, but also for global MPXV surveillance, as lineages circulating within Africa have the potential to contribute to regional or international spread.

MPXV is phylogenetically classified into two main clades: Clade I (previously the Congo Basin clade) and Clade II (formerly the West African clade). Historically, Clade I, predominantly found in Central Africa, is linked to more severe illness and higher mortality. In contrast, Clade II is associated with milder disease and lower-case fatality rates. Both Clade I and Clade II are further divided into subclades a and b [9]. Early reports from the 2024/2025 outbreak in Uganda indicate the predominance of Clade Ib, although the possibility of multiple introductions and regional evolution necessitates detailed genomic characterization. A particularly concerning feature is the observation of accelerated microevolution in some MPXV lineages, which has been associated with host APOBEC3 cytidine deaminase activity [10], [11]. These mutations may affect viral fitness, transmissibility, and even diagnostic assay performance if they occur in primer/probe binding sites.

The 2024/2025 Uganda MPXV outbreak was characterized by geographically widespread transmission, increased case detection through enhanced surveillance, and integration of genomic sequencing into routine outbreak response. The scale and duration of the outbreak provided a unique opportunity to investigate MPXV genomic diversity in real time and to examine how viral lineages evolved and disseminated across different regions of the country.

Genomic epidemiology has become an essential tool for outbreak investigation, enabling high-resolution reconstruction of transmission chains, identification of introduction events, and monitoring of viral evolution [12], [13]. For MPXV, genome sequencing has revealed lineage-specific patterns of spread, accumulation of mutations potentially driven by host-mediated editing mechanisms, and evidence of micro-evolution during sustained transmission [10], [14]. However, much of the high-resolution genomic insight into MPXV evolution and transmission has emerged from outbreaks outside Africa, where sequencing intensity and data availability have been greatest, underscoring the need for locally generated genomic data to capture region-specific evolutionary dynamics.

Understanding the evolutionary dynamics of MPXV during outbreaks is particularly important for countries with historically endemic transmission, where repeated zoonotic spillover events may intersect with human-to-human transmission. Genomic analyses can help distinguish between multiple introductions and sustained transmission, assess genetic diversification over time, and identify signatures of adaptation that may influence transmissibility, virulence, or diagnostic performance. Such insights are essential for informing public health interventions, refining surveillance strategies, and strengthening preparedness for future outbreaks [12], [13].

Here, we present a comprehensive genomic characterization of MPXV isolates from the 2024/2025 outbreak in Uganda. By integrating whole-genome sequencing with phylogenetic analysis and global reference datasets, we elucidate the genetic diversity and lineage structure of the circulating virus, providing critical insights into the evolutionary dynamics specific to this outbreak.

## Methods

### Ethics statement

This work adhered to the principles of the Declaration of Helsinki. The Uganda National Health Laboratory Services Research and Ethics Committee approved the protocol (UNHLS-2025-133) and waived the requirement for individual informed consent, acknowledging that sampling was performed for active surveillance as part of the national Mpox outbreak response in Uganda.

### Sample collection and sequencing

During the MPXV outbreak declared in Uganda in July 2024, a random subset of symptomatic suspected cases that tested positive for MPXV by real-time PCR were included in this study. Samples were collected as part of the public health emergency response from affected individuals across 44 districts in Uganda and included blood, genital swabs, lesion swabs, oropharyngeal swabs, rash swabs, saliva, and semen.

DNA was extracted using QIAamp DNA Kit (Qiagen, Hilden, Germany) and screened using the CDC Non-Variola Orthopoxvirus Real-Time PCR Primer and Probe Set (CDC, EUA) [15] at the Central Emergency Response and Surveillance Laboratory (CERSL), Ministry of Health, Uganda. PCR amplification was performed on a CFX96 Real-Time PCR Detection System (Bio-Rad Laboratories, Inc., USA), with an initial denaturation at 95 °C for 8 minutes, followed by 40 cycles consisting of denaturation at 95 °C for 5 seconds and annealing/extension at 60 °C for 30 seconds. Cycle threshold (Ct) values were assessed for quality control using the CFX96 software. Extracted DNA was then subjected to tiled amplicon amplification using the Illumina Microbial Amplicon Prep (iMAP) Kit in combination with the Yale Mpox primer scheme.

For Illumina sequencing, amplicons generated using the iMAP workflow were processed following the remainder of the library preparation protocol, including indexing and library clean-up steps as specified by the manufacturer. The prepared libraries were quantified, pooled, and sequenced on the Illumina MiSeq platform (Illumina Inc., San Diego, CA, USA).

For nanopore sequencing, the amplified products were prepared using the Oxford Nanopore Rapid Barcoding Kit 24 V14 following the manufacturer’s instructions. Barcoded libraries were pooled and sequenced on an R10 Flow Cell using the Oxford Nanopore GridION platform (Oxford Nanopore Technologies [ONT], Oxford, UK).

### Bioinformatics analysis

Raw FASTQ reads were processed using the ARTIC (version 1.4.4) Nextflow pipeline to generate consensus MPXV genomes [16]. Generated consensus sequences were subsequently assigned to clades using Nextclade [17]. Additional Clade Ib MPXV genomes and associated metadata were retrieved from Pathoplexus (https://doi.org/10.62599/PP_SS_496.1), GISAID (https://doi.org/10.55876/gis8.260107gz), and NCBI Virus databases [18]–[22]. To avoid redundancy, genomes present in both Pathoplexus and NCBI were identified and removed.

Multiple sequence alignment was performed using Squirrel, with the quality-control (QC) mode enabled to flag single nucleotide polymorphisms (SNPs) located near regions of ambiguous bases (N), clusters of unique SNPs, reversions to reference alleles, and convergent mutations [23]. Sites flagged during QC were subsequently used for maximum-likelihood phylogenetic reconstruction within Squirrel.

The apobec3-phylo mode in Squirrel was additionally enabled to characterize APOBEC3-like and non-APOBEC3 mutation patterns across the phylogeny in a descriptive manner. The phylogenetic tree was rooted using the historical MPXV genome KJ642613|human|DRC|Equateur|1970-09-01 as the outgroup. The phylogenetic tree was visualized using the interactive tree of life (iTOL) [24]

## Results

### Demographics

We analyzed 511 PCR-confirmed MPXV genomes from 44 districts across Uganda (Fig 1), all meeting the inclusion criterion of ≥70% genome coverage. Among the cases associated with these samples, 262 (51.3%) were male and 214 (41.9%) were female, while sex was unrecorded for 35 (6.8%) individuals. The mean patient age was 27 years (range: 1–60 years). To provide regional context, we additionally incorporated 895 publicly available clade Ib MPXV genomes with ≥70% coverage collected between 2023 and 2025. These were retrieved from GISAID (n = 590), Pathoplexus (n = 303), and NCBI (n = 2), with the majority originating from Uganda and the Democratic Republic of Congo (Table 1).

**Table 1.** Country-level distribution of publicly available clade Ib MPXV genomes (≥70% genome coverage) from GISAID, Pathoplexus, and NCBI.

| Country | Total (n) |
| --- | --- |
| DRC | 556 |
| Uganda | 143 |
| Burundi | 92 |
| Germany | 18 |
| USA | 16 |
| Canada | 10 |
| Congo | 10 |
| Thailand | 10 |
| Malawi | 6 |
| United Kingdom | 5 |
| China | 3 |
| India | 3 |
| Kenya | 3 |
| Netherlands | 3 |
| Pakistan | 3 |
| Ireland | 2 |
| Italy | 2 |
| Oman | 2 |
| Sweden | 2 |
| Portugal | 2 |
| Australia | 1 |
| Brazil | 1 |
| Japan | 1 |
| Rwanda | 1 |

**Figure 1.**
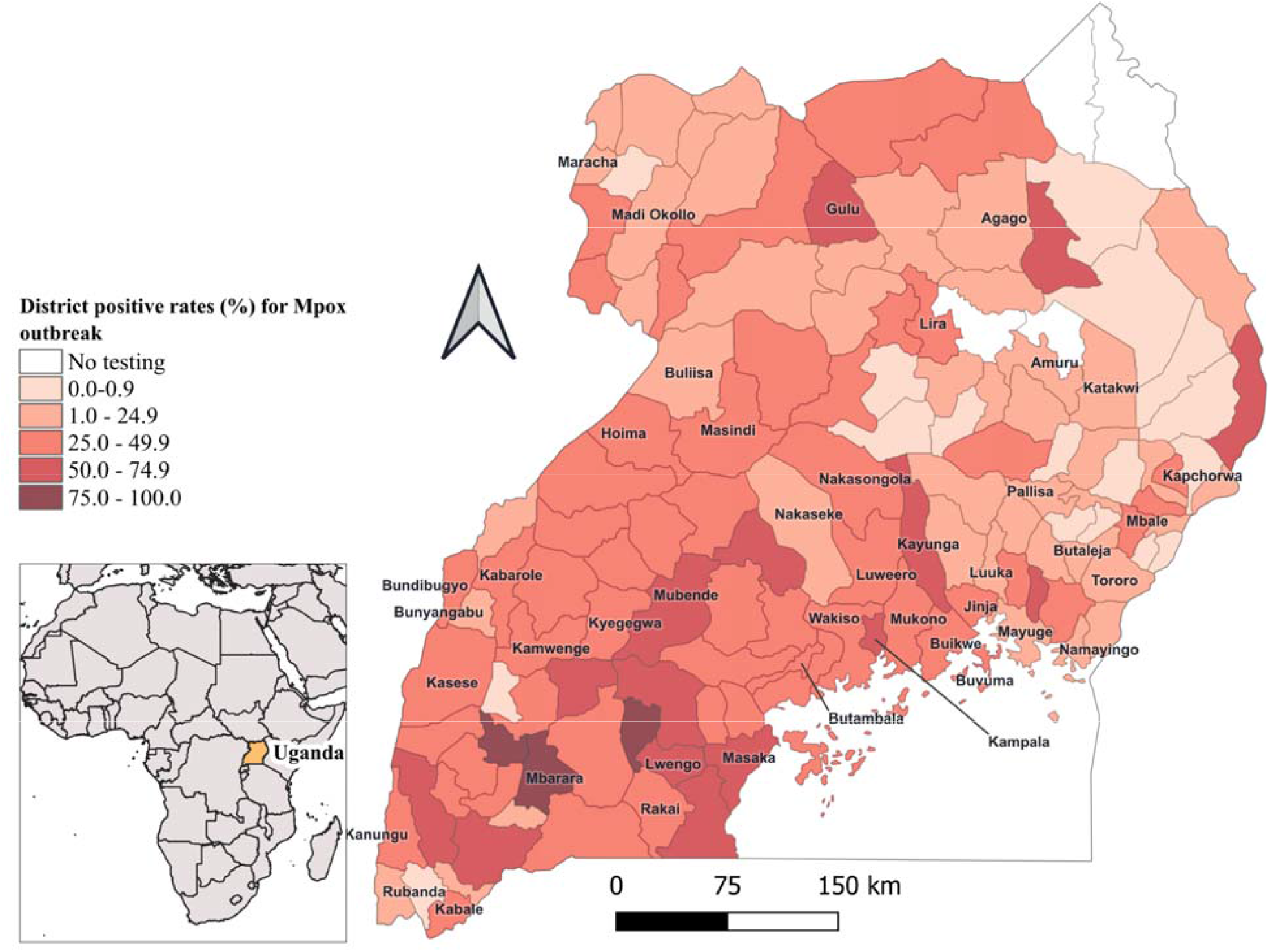
Geographic distribution of sample collection sites across Uganda. Map of Uganda showing district-level MPXV PCR positivity rates based on national surveillance data collected between July 2024 and December 2025. The map was generated using QGIS (version QGIS-OSGeo4W-3.44.6-1; https://qgis.org/).

### Phylogenetic Structure and Phylogeographic Dynamics

The analyzed sequences exhibited genomic signatures indicative of sustained human-to-human transmission, consistent with ongoing local spread. Phylogenetic reconstruction resolved the Clade Ib sequences into two primary clusters (Clusters 1 and 2), each subdivided into two distinct subclusters. Cluster 1 was predominantly composed of sequences from the Democratic Republic of the Congo (DRC) and displayed comparatively lower genetic diversity. In contrast, Cluster 2 exhibited greater genetic diversity and a broader geographic distribution. Notably, the majority of Ugandan sequences (n =574) grouped within Subcluster 2 of Cluster 2, which represented the most genetically diverse lineage in the dataset. However, a minority of Ugandan isolates were also identified in Subcluster 1 of Cluster 2 (n = 9) and Subcluster 2 of Cluster 1 (n = 64), confirming the co-circulation of multiple lineages. Phylogeographic analysis further clarified these patterns, identifying multiple independent cross-border introductions into Uganda and highlighting the critical role of regional connectivity in driving the observed transmission dynamics (Fig 3).

**Figure 3.**
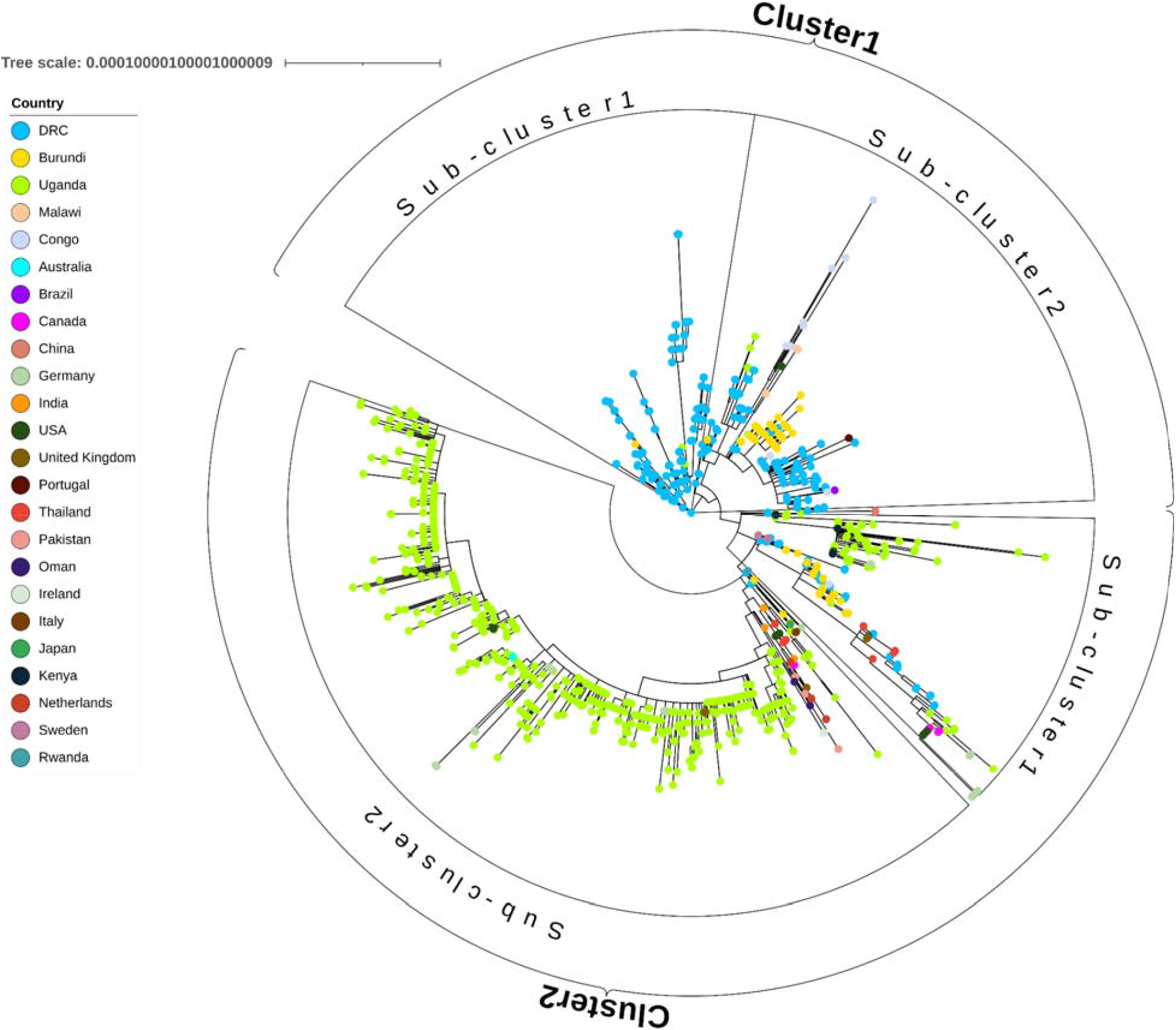
Maximum-likelihood phylogeny of clade Ib MPXV genomes from Uganda and public datasets (≥70% genome coverage), showing two major clusters. Clusters and subclusters 1/2 are study-defined groups.

## Discussion

This work presents a comprehensive genomic characterization of the clade Ib mpox outbreak in Uganda, synthesizing locally generated genomes with a broad dataset of regionally available sequences. By implementing a strict quality threshold of ≥70% genome coverage, we ensured robust phylogenetic inference while maintaining spatial and temporal representativeness.

The phylogenetic analysis resolved two major clusters within clade Ib, each subdivided into distinct subclusters, reflecting substantial lineage diversification. The predominance of Ugandan sequences within the highly diverse Cluster 2 (Subcluster 2) is indicative of sustained local transmission following successful introduction events, rather than repeated, short-lived spillovers. High genetic diversity within this subcluster is consistent with prolonged circulation and onward transmission within Uganda. Conversely, Cluster 1 was dominated by sequences from the Democratic Republic of Congo (DRC) and displayed lower genetic diversity. While this could signal a more established, stable transmission network in the DRC given the longstanding endemicity of the virus, such interpretations warrant caution due to potential sampling biases.

Phylogeographic reconstruction provides compelling evidence of multiple cross-border introductions into Uganda, highlighting the critical role of regional connectivity in shaping outbreak dynamics. Population mobility, porous borders, and frequent cross-border trade between Uganda and neighboring countries, particularly the DRC, appear to facilitate repeated viral introductions. Importantly, these introductions have seeded local transmission chains rather than remaining epidemiological dead ends, as evidenced by the clustering patterns and sustained human-to-human transmission signatures.

A significant limitation of this analysis is the presence of sampling disparities. The dataset is heavily skewed toward Uganda and the DRC, reflecting differences in sequencing capacity and surveillance intensity across the region. Consequently, the observed phylogenetic structure may obscure unsampled diversity or alternative transmission routes in under-represented neighboring countries. Despite this spatial bias, the data strongly supports the conclusion that cross-border mobility is a primary driver of the outbreak.

Overall, these findings underscore the urgency of harmonized cross-border data sharing and the strengthening of regional sequencing capacity. Enhanced surveillance is essential to mitigate the effects of sampling disparities and to support the early detection of lineage expansion across East and Central Africa.

## Data availability

The genomic fasta sequences generated during this study are available at Pathoplexus (https://doi.org/10.62599/PP_SS_925.1).

The findings of this study are also based on publicly available sequence data and metadata associated with https://doi.org/10.62599/PP_SS_496.1 and https://doi.org/10.55876/gis8.260107gz.

## Author contributions

Conceptualization: S.K., A.A., I.S., S.N., D.J., G.M., D.P.K, A.S., S.K.T., I.S. Formal analysis: S.K., A.A., J.M.K., C.M., I.S., G.M. Funding acquisition: S.N., I.S., S.T.K. Methodology: S.K., A.A., J.M.K., H.R.O., J.S., W.T., G.T., S.W., M.M., M.N., V.Z.N., C.M., I.S., G.M., A.S, I.M, B.L, E.M.R, M.O, S.G. Validation: S.K., A.A., B.A.K., H.O., C.T., N.A., V.Z.N., N.A., G.M., A.S., S.K.T., I.S. Visualization: S.K., I.S., J.M.K. Software: S.K., J.M.K., I.S., G.M. Writing – original draft: S.K., A.A., B.A.K., V.Z.N., I.S., H.O., C.T., N.A., S.N., D.J., G.M., A.A., A.S., S.K.T., I.S. Writing – review and editing: All authors.

## Acknowledgement

We acknowledge the Ministry of Health Uganda and the Central Public Health Laboratories (CPHL) for supporting sample collection, diagnostic testing, and genomic sequencing during the Mpox outbreak response. We are grateful to the Genomics Core Facility at CPHL for sequencing support and to the field epidemiology and surveillance teams for their commitment to timely case investigation and sample coordination.

We also gratefully acknowledge all data contributors and their Originating laboratories responsible for obtaining the specimens, and their submitting laboratories for generating the genetic sequence and metadata and sharing via the GISAID Initiative, Pathoplexus, NCBI virus, on which this research is partly based.

